# Clinical, environmental, and sociodemographic factors in ethnic differences in incidence of type 2 diabetes complications and mortality in a Dutch dynamic prospective primary care cohort: a DIAMANT study

**DOI:** 10.64898/2026.08.12.26360278

**Authors:** M. Muilwijk, B.T. Strooij, P.J.E.M. Elders, F. Rutters, G. Nijpels, I. Vaartjes, J.A. Overbeek, R.M.C. Herings, J. Lakerveld, M.T. Blom, J.W.J. Beulens

## Abstract

**Introduction:** Ethnic minority populations are disproportionately affected by type 2 diabetes (T2D). We investigated ethnic differences in the risks of diabetes-related complications and mortality in the Netherlands, and identified clinical, sociodemographic and environmental determinants associated with these differences.

**Methods:** We included 175,112 adults with T2D from the dynamic prospective primary care cohort DIAMANT. DIAMANT data were linked to national registries from Statistics Netherlands and GECCO, a database integrating geographic, environmental and contextual exposures. Ethnic differences in complications risks were estimated using Cox proportional hazards models. Potential mediating factors were explored using machine-learning-based variable selection and association decomposition approaches.

**Results:** At baseline, mean age was 65.4 (SD 12.3) years, 46.6% were women and median T2D duration was 11.3 [IQR 7.2; 15.8] years. Substantial heterogeneity in complication risk was observed across ethnic groups compared with Dutch-origin individuals. Retinopathy risk was consistently higher across nearly all non-Dutch groups (HRs 1.37-2.37). For macrovascular complications, elevated risks were mainly observed among Surinamese and Turkish individuals, including heart failure (HR 1.30 and 1.46, respectively). In contrast, individuals of Indonesian and Moroccan origin showed similar or lower risk for most complications. Environmental exposures (e.g. air pollution, temperature) and sociodemographic factors (e.g. main benefit, household composition) accounted for a substantial attenuation of several observed associations.

**Discussion:** Substantial ethnic differences exist in risks of T2D complications and mortality, which showed to be heterogeneous across outcomes and populations. Our findings suggest that a considerable proportion of these disparities is attributable to differences in environmental and sociodemographic context, highlighting the importance of interventions that take into account differences in environmental and socio-demographic context.

## Introduction

Ethnic minority populations in Europe are disproportionately affected by type 2 diabetes (T2D), with prevalence estimates two to six times higher than among populations of European origin [1-3]. T2D is characterized by elevated blood glucose levels, which lead to both macrovascular and microvascular complications [4]. Across European settings, ethnic minority populations with T2D show heterogeneous risks of these complications compared with their European counterparts, with risk patterns varying by type of complication and host country [5-7]. For instance, South Asian populations consistently exhibit higher microvascular risk, whereas African-origin populations display macrovascular risks in some contexts. Prospective studies in Europe, primarily from the United Kingdom, likewise report higher microvascular complication risks across ethnic groups, while macrovascular risks differ more substantially between groups when compared to White Europeans [3, 8-10].

Although several studies in the Netherlands have examined ethnic differences in T2D complications, the evidence base remains limited. Many studies are over 15 years old, include modest sample sizes or assess only a subset of complications, and may therefore not fully reflect current patterns in a changing and increasingly diverse population. Earlier studies showed higher nephropathy risk among South-Asian Surinamese compared with Dutch individuals [11, 12], and differential ischemic heart disease risk among Turkish and South-Asian Surinamese populations [13]. More recent cross-sectional analyses from the Dutch HELIUS cohort indicate increased nephropathy and coronary heart disease among most ethnic minority populations compared to Dutch-origin, though not uniformly across all groups [6]. Psychological complications also differ, with stronger associations between T2D and depression reported among individuals of non-Dutch descent [14].

Ethnic differences in T2D complications may arise from unequal distributions of sociodemographic factors, health-related behaviors, environmental exposures and T2D management, as well as genetic and physiological determinants that influence susceptibility [15, 16]. Socioeconomic disadvantage, more common in several minority populations, is linked to poorer glycemic control and elevated complication risks [17]. Additionally, ethnic minority populations more frequently reside in neighborhoods with higher exposure to environmental stressors, including air pollution and noise [18, 19], which have been associated with adverse T2D outcomes [20]. Together, these interrelated determinants underscore the need for a comprehensive examination of the mechanisms underlying ethnic differences in T2D complications, with explicit attention to the joint contribution of clinical, contextual and environmental factors.

Therefore, this study aims to assess differences in the risks of T2D complications and mortality among ethnic minority groups living in the Netherlands compared with the Dutch-origin population, and to evaluate the extent to which sociodemographic, clinical and environmental factors explain these differences. To this end, we use an integrated analytical approach combining machine learning with traditional epidemiological methods.

## Methods

### Population and study design

We used data from the DIAMANT cohort, a dynamic, prospective registry of people with diabetes managed in primary care in the Netherlands [21]. DIAMANT contains anonymized electronic health records from approximately 20% of Dutch general practices, with data available from 2004 onwards, extracted via the PHARMO GP Database with consent from participating general practitioners. The cohort is maintained by “Stichting Informatievoorziening voor Zorg en Onderzoek” (STIZON) and includes diagnoses, symptoms, laboratory measurements and prescriptions. Demographic characteristics and primary care diagnoses in DIAMANT are representative of the Dutch population with T2D[21]. DIAMANT data from 2015-2019 were pseudonymized at the source and linked to nationwide administrative registries from Statistics Netherlands using probabilistic linkage based on birthdate, registered sex and postal code. All analyses were conducted within the secure remote environment of Statistics Netherlands under strict privacy conditions.

We included adults with a diagnosis of T2D (ICPC T90.02) or at least two prescriptions of glucose-lowering medication (ATC A10B) within six months. Cohort entry was defined as the latest of: 1) 1 January 2015, 2) the start of registration in the general practice, or 3) the first recorded T2D diagnosis. Thus, individuals entered the cohort only once they were both registered in a participating general practice and had evidence of T2D. Individuals residing in institutional or unknown households and those receiving specialist-led care were excluded. For each outcome, individuals with a prior diagnosis were excluded to ensure incident cases. Participants were followed from the cohort entry date until the earliest of: 1) first occurrence of the outcome, 2) end of registration in a participating general practice, 3) death, or 4) 31 December 2019. Follow-up ended when individuals were no longer registered in a participating general practice. The study followed STROBE reporting guidelines [22] and was approved by the DIAMANT cohort governance board. The Medical Ethical Committee of Amsterdam UMC (2022.0612) waived ethical approval because pseudonymized routine care data were used. Patients were informed by their general practitioner that their data could be used for scientific research and could object.

### Variables

#### Ethnicity

Ethnicity was operationalized according to the Statistics Netherlands’ definition of migration background, based on individuals’ and parents’ country of birth [23]. For first-generation migrants, the ethnic group was defined by the person’s country of birth; for second-generation migrants, by the parent’s country of birth (mother’s origin used in case of discordance). Country of birth is a validated proxy for distinguishing population groups in the Netherlands [24]. Groups were classified according to the new population-origin taxonomy of Statistics Netherlands into: Morocco, Turkey, Suriname, Dutch Caribbean, Indonesia, other European (excluding the Netherlands), and non-European (excluding the above) [23]. “Dutch-origin” refers to individuals born in the Netherlands with two Dutch-born parents.

#### Outcomes

We examined microvascular, macrovascular, psychological and mortality outcomes. Diagnoses were identified using ICPC codes from primary care records and cause-specific mortality was identified using ICD codes from Statistics Netherlands national death registry.

Microvascular complications included retinopathy (ICPC F83 series), nephropathy (U99.01), neuropathy (N94.02) and foot ulcer (S97). Macrovascular complications comprised angina pectoris (K74), myocardial infarction (K75), other ischemic diseases (K76), transient ischemic attack (K89), cerebral vascular accident (K90 series), peripheral artery disease (K92.01) and heart failure (K77 series). Depression was defined using ICPC P76. All-cause mortality was based on ICPC A96-codes, and cardiovascular mortality by ICD codes I05 to I78. Composite outcomes were defined as any microvascular complication, or any macrovascular complication.

#### Covariates

Age, sex, T2D duration and baseline HbA1c were determined at index date. Follow-up time was defined as described above. As HbA1c had 23.3% missingness, it was imputed using multiple imputation with the MICE package in R. No other covariates had missing data.

#### Explanatory variables

Potential explanatory variables (full list, detailed definitions and exposure assessment methods in Supplementary File 1) were categorized as:

1. health indicators at baseline, e.g. BMI and Charlson co-morbidity index;
2. medication use, including polypharmacy and Anatomical Therapeutic Chemical (ATC) classification levels 3, 4, 5 and 7, with higher levels providing progressively more specific detail about a medication;
3. healthcare use, e.g. participation in diabetes care program and annually measured kidney function;
4. individual-level socio-economic status, e.g. disposable household income and household composition;
5. lifestyle, e.g. smoking, healthy diet advice received;
6. environmental exposures, e.g. air pollution, noise, greenspace.

Clinical and lifestyle variables were derived from DIAMANT. Socioeconomic indicators were obtained from Statistics Netherlands. Environmental exposures were obtained from the Geoscience and Health Cohort Consortium (GECCO), the Dutch Health Monitor and Statistics Netherlands. GECCO provides longitudinal environmental data at various geospatial levels in the Netherlands [25]. Address-level exposure data obtained from GECCO were aggregated to mean values of 6-digit postal code (PC6) areas, representing small areas that can be crossed on foot in about 8 minutes. The health monitor periodically maps the health, well-being and lifestyle of the Dutch population, using data collected by Statistics Netherlands in collaboration with municipal health services (GGD-en) and the National Institute for Public Health and the Environment (RIVM) via online and paper questionnaires. A stratified sampling method ensures a representative sample of residents per neighborhood across the Netherlands.

### Statistics

Ethnic differences in T2D complications were assessed using Cox proportional hazards models, and the proportional hazards assumption was verified using Schoenfeld residuals. To protect confidentiality, results were suppressed when <10 events occurred.

To identify variables most strongly associated with ethnic differences in each complication, we used gradient-boosted decision trees (XGBoost) [26]. This machine-learning approach was used to flexibly handle multiple correlated exposures and potential non-linear associations [27, 28]. Subsequent analyses using Cox proportional hazard models allowed for transparent estimation and interpretation of effect estimates [22]. Variable importance was derived using Shapley Additive Explanation (SHAP) values [29], which quantify each variable’s marginal contribution to model predictions. For each outcome, the 20 top-ranked variables by SHAP were selected. Highly correlated variables (Spearman’s ρ ≥0.9) were reduced by retaining the variable with the highest SHAP value.

To estimate the extent to which selected variables mediated ethnic differences, we applied the difference method within Cox models. We first estimated the total effect of ethnicity, then re-estimated hazard ratios after adding the selected explanatory variables (direct effect). The indirect effect was calculated as the percentage reduction in log-HR. Statistical significance of mediation was evaluated using a permutation test (n=1,000) in which ethnicity was randomly permuted to obtain empirical p-values [30].

To assess whether patterns differed by migration generation, we conducted exploratory analyses stratified into first- and second-generation individuals. Because second-generation migrants are generally younger than first-generation migrants and thus have lower underlying risks of T2D complications, these analyses were performed for descriptive purposes only and not intended as causal comparisons. Cox proportional hazards models identical to the primary models were re-estimated within each generation stratum.

## Results

### Cohort and baseline characteristics

A total of 249,337 out of 286,652 individuals (87.0%) with T2D from the DIAMANT cohort were successfully linked to the Statistics Netherlands databases (Figure 1). After application of the inclusion criteria, 175,112 individuals were included in the analysis. The percentage of women ranged from 44.8% among non-Europeans, to 59.3% among Dutch Caribbeans (Table 1). The mean age at baseline was lowest among non-Europeans (55.5 years [SD 11.4]), and highest among Dutch-origin (66.6 years [SD 11.9]). Median follow-up duration was 5.0 years [IQR 3.8; 5.0].

**Table 1.** Baseline Characteristics Stratified By Ethnicity.

|  | Dutch-origin<br>(N=139,110) | Indonesian<br>(N=4,972) | Moroccan<br>(N=3,636) | Surinamese<br>(N=5,480) | Turkish (N=3,725) | Dutch Caribbean<br>(N=1,326) | European<br>(N=10,306) | non-European<br>(N=6,557) |
| --- | --- | --- | --- | --- | --- | --- | --- | --- |
| **Sex, Women (N; %)** | 64,185 (46.1%) | 2,280 (45.9%) | 1,753 (48.2%) | 2,922 (53.3%) | 1,809 (48.6%) | 786 (59.3%) | 4,851 (47.1%) | 2,935 (44.8%) |
| **Age (years)** | 66.6 (11.9) | 64.0 (11.3) | 57.8 (12.3) | 57.7 (11.0) | 56.6 (11.7) | 58.6 (11.4) | 66.4 (12.1) | 55.5 (11.4) |
| **Diabetes duration (years)** | 11.4 [7.29; 15.8] | 11.7 [7.65; 16.0] | 12.0 [7.71; 16.5] | 12.4 [7.96; 16.8] | 11.2 [7.37; 15.2] | 11.1 [6.89; 15.2] | 11.0 [6.84; 15.6] | 9.34 [5.49; 14.0] |
| **HbA1c (mmol/mol)** | 50.0 [44.1; 58.0] | 51.0 [45.0; 58.0] | 53.0 [47.0; 63.0] | 51.0 [46.0; 60.0] | 54.0 [47.0; 65.0] | 53.0 [47.0; 64.0] | 50.0 [44.0; 58.0] | 52.0 [46.0; 62.0] |
| Missing (N; %) | 31,720 (22.8%) | 1,228 (24.7%) | 822 (22.6%) | 1,443 (26.3%) | 1,030 (27.7%) | 353 (26.6%) | 2,554 (24.8%) | 1,602 (24.4%) |
| **Follow-up time (years)** | 5.00 [3.79; 5.00] | 5.00 [4.14; 5.00] | 5.00 [4.49; 5.00] | 5.00 [4.47; 5.00] | 5.00 [4.03; 5.00] | 5.00 [3.77; 5.00] | 5.00 [3.64; 5.00] | 5.00 [3.36; 5.00] |

*Baseline characteristics are shown by percentages and numbers (N) for categorical variables, by means and standard deviations for normally distributed numerical variables, and by medians and interquartile ranges for not-normally distributed numerical variables.*

**Figure 1.**
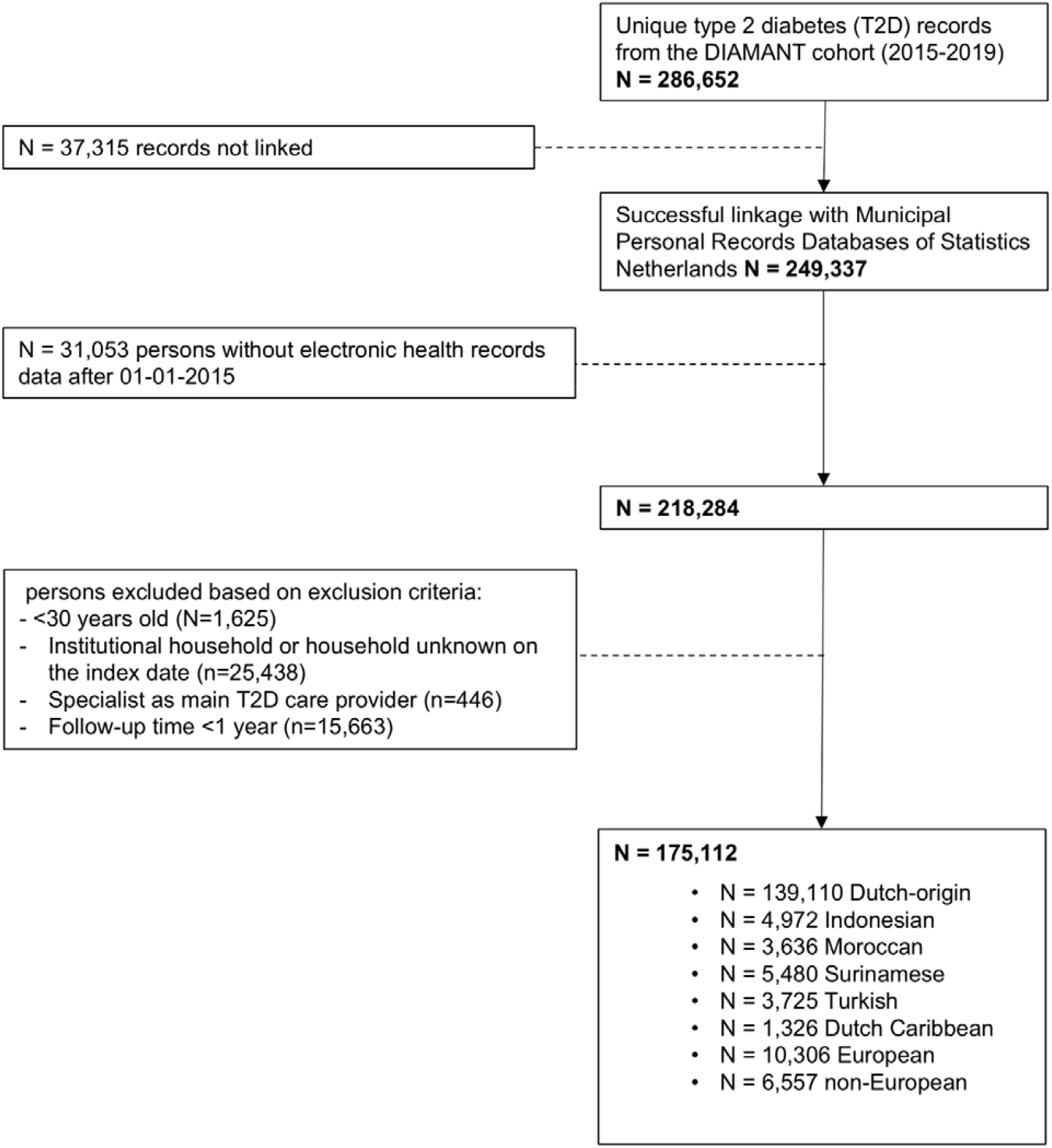
Flowchart on inclusion of people with type 2 diabetes into the study

### Selected potential mediators

Across complications, SHAP-based ranking identified a consistent set of influential predictors, spanning all considered domains (Supplementary File 2). The reported n indicates the number of models in which a variable ranked among the top 20 predictors. Baseline health predictors most frequently ranking among the top 20 predictors included the Charlson comorbidity index (n=14), baseline blood pressure (n=9), and baseline BMI (n=7). Environmental exposures were consistently represented by environmental temperature (n=12), air pollution NH_3_ (n=11) and air pollution SO_2_ (n=11). Healthcare use included annual measured kidney function testing (n=10) and LDL cholesterol testing (n=8), and participation in structured diabetes care programs (n=7). Lifestyle factors included baseline smoking status (n=13) and alcohol consumption (n=4). SES was reflected by household type (n=10), household composition (n=9) and primary source of benefits (n=7). Medication-related predictors included use of glucose-lowering drugs (ATC A10 and subcategories; n=9) and antithrombotic agents (B01 and subcategories; n=6).

### Ethnic differences in mortality

In age- and sex-adjusted models, all-cause mortality differed across ethnic groups compared with individuals of Dutch-origin. Lower risks were observed among Moroccans (HR 0.78 [95%-CI 0.62; 0.98]) and non-Europeans, whereas higher risks were seen among Turks (HR 1.26 [95%-CI 1.03; 1.55]) (Figure 2; Supplementary File 3-4). After additional adjustment for HbA1c and T2D duration, estimates were only minimally attenuated, but no longer statistically significant among Moroccans, while non-Europeans continued to show lower risks for all-cause mortality (HR 0.79 [95%-CI 0.64; 0.96]). No statistically significant differences in all-cause mortality were observed among Indonesians, Surinamese, Dutch Caribbeans or Europeans. For CVD mortality, lower risks were consistently observed among Moroccans and non-Europeans across models, including in the fully adjusted model (e.g. HR 0.61 [95%-CI 0.45; 0.82] for non-Europeans). In mediation analyses, the higher all-cause mortality risk observed in Turkish individuals was largely explained by environmental exposures. Adjustment for these factors attenuated the excess risk substantially and shifted the estimate below the reference level, indicating that environmental differences more than accounted for the initially observed risk difference. Environmental factors contributed most strongly to this attenuation (119 and 115%), followed by SES (37.8% and 36.9%), and healthcare use (34.8% and 42.3%). Medication use and lifestyle factors (including antithrombotic agents and baseline smoking) explained a smaller proportion of the lower risk among non-Europeans, ranging from 10.3-17.9% (Figure 3-4; Supplementary Files 5-6).

**Figure 2.**
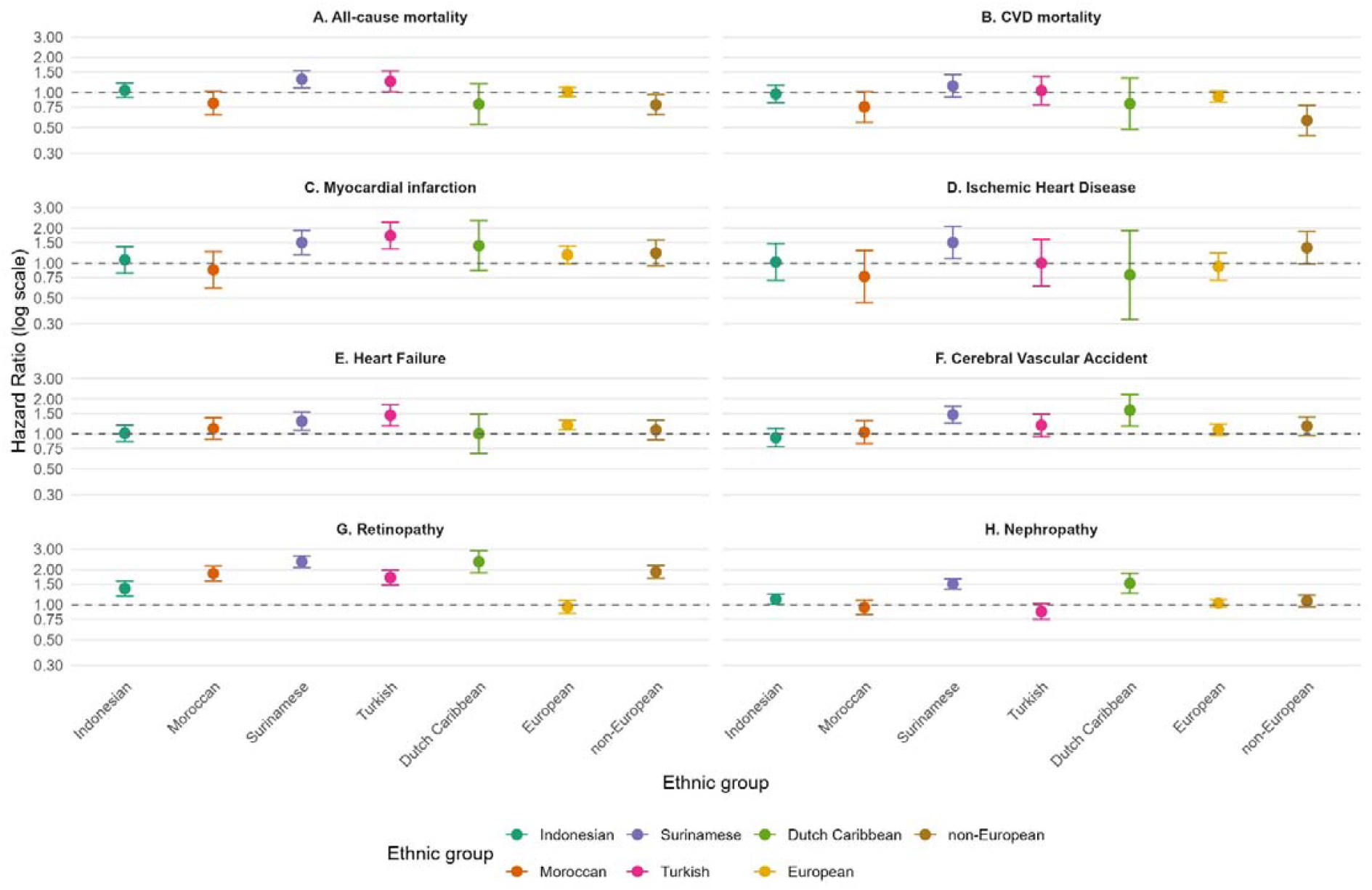
Forest plots of associations between ethnicity and type 2 diabetes complications. Dutch was set as reference category (HR=1.0).

**Figure 3.**
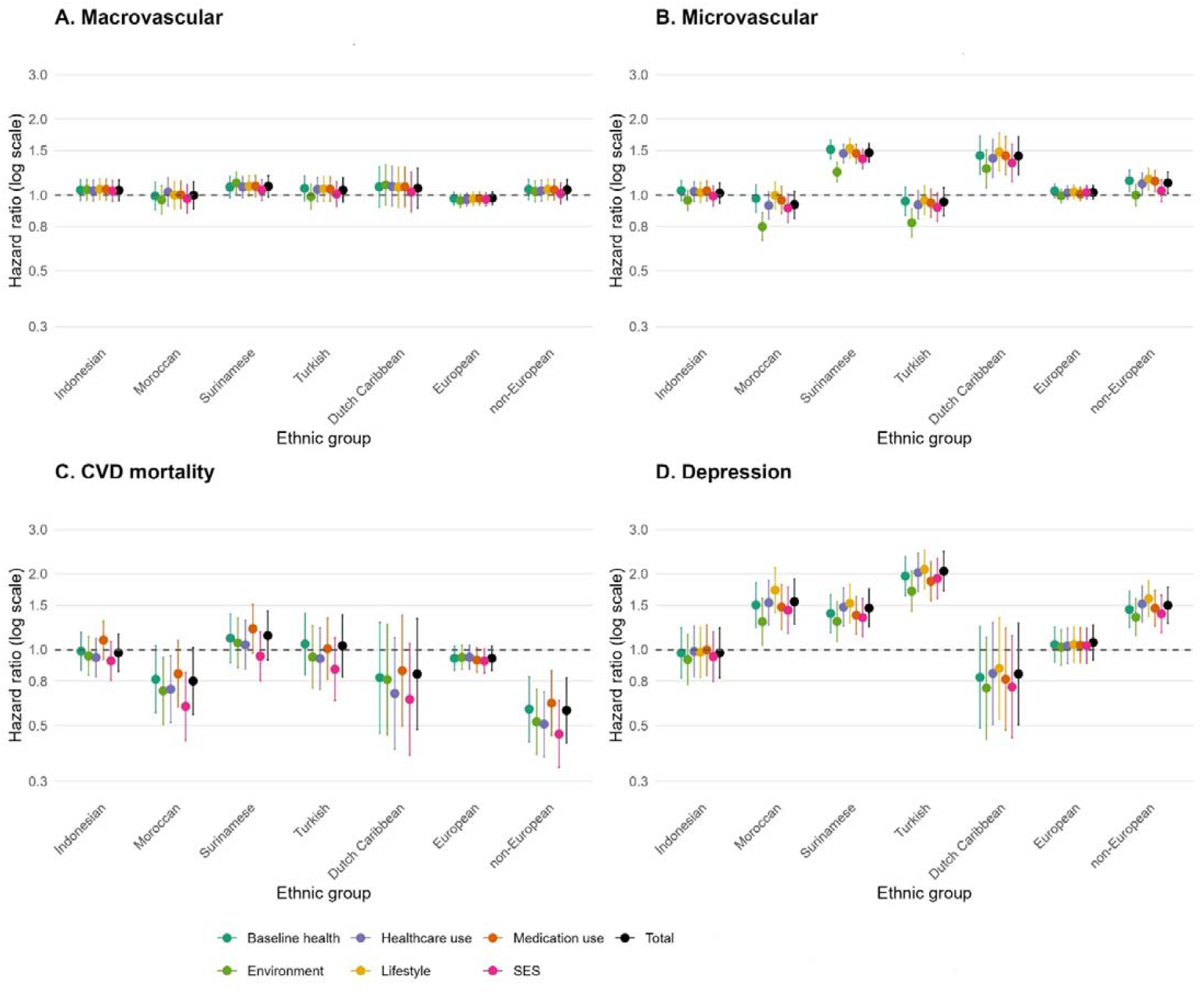
Associations between ethnicity and type 2 diabetes complications with different sets of mediators. Dutch was set as reference category (HR=1.0). The ‘Total’ (in black) estimate, is the estimated hazard ratio without addition of mediator sets, while the colors represent estimates with adjustment for the different sets of mediators. Dutch was set as reference category. The ‘Total’ (in black) estimate, is the estimated hazard ratio without addition of mediator sets, while the colors represent estimates with adjustment for the different sets of mediators. Included factors in each mediator set are listed hereafter. A) Macrovascular complications. 1) Baseline health: Charlson Comorbidity Index; baseline LDL; baseline kidney function; baseline blood pressure; baseline BMI; baseline kidney function; baseline HDL. 2) Healthcare use: participation in a diabetes care program; annually measured blood pressure; annually measured BMI. 3) Medication use: Atorvastatin (C10AA05). 4) Environment: environmental temperature; air pollution SO_2_; compliance with physical activity norm at the neighborhood level; air pollution NH_3_. 5) Lifestyle: baseline smoking. 6 SES: household composition; population density; household type; home ownership. B) Microvascular complications. 1) Baseline health: Charlson Comorbidity Index; baseline triglycerides; baseline LDL; baseline blood pressure. 2) Healthcare use: annually measured kidney function; annual control of the foot, participation in a diabetes care programme. 3) Medication use: antithrombotic agents (B01A); drugs used in diabetes (A10); calcium channel blockers (C08); insulin and analogues (A10A). 4) Environment: population density; environmental temperature; turnout elections. 5) Lifestyle: baseline smoking. 6 SES: household composition, household type, main benefit. C) CVD Mortality. 1) Baseline health: baseline BMI; Charlson Comorbidity Index; baseline kidney function . 2) Healthcare use: participation in a diabetes care program; annually measured LDL; annually measured HbA1c; annually measured kidney function; annual control of the foot; annually measured blood pressure. 3) Medication use: antithrombotic agents (B01); sulfonamides plain (C03CA); vitamin K agonists (B01AA). 4) Environment: air pollution SO_2_; environmental temperature; compliance with physical activity norm at the neighborhood level; the percentage widowed households at the neighborhood level. 5) Lifestyle: N.A. 6 SES: household type; household composition; main benefit; home ownership. D) Depression. 1) Baseline health: Charlson Comorbidity Index; baseline blood pressure, fatigue/weakness; insomnia/other sleep disorder (icpc P06). 2) Healthcare use: annually measured kidney function; annually measured LDL; annually measured BMI . 3) Medication use: analgesics (NO2). 4) Environment: air pollution NH3; environmental temperature; landuse mix 1000z; air pollution SO2; density fast food restaurants 500m; % households with a low income in neighborhood; schools within 3km; % rental homes neighborhood. 5) Lifestyle: baseline smoking. 6 SES: main benefit; household type; household composition.

**Figure 4.**
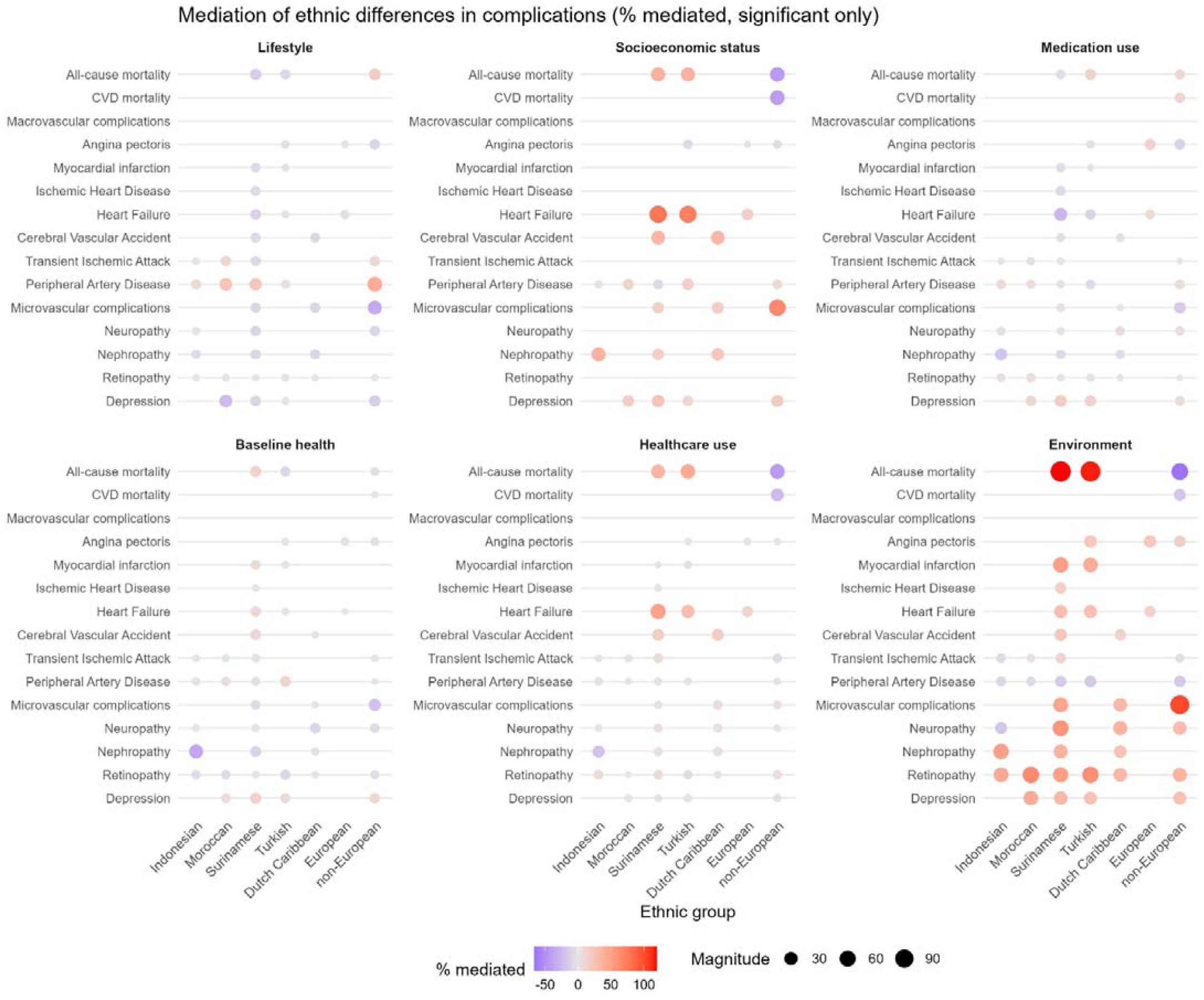
Percentage mediated by different mediator sets for type 2 diabetes complications with a statistical significant ethnic difference compared to Dutch-origin. Colors represent the magnitude and direction of mediation (blue = negative, red = positive, grey = around zero), and point size reflects the absolute magnitude of the mediated proportion. Only outcomes with a statistically significant ethnic difference (P < 0.05) are shown.

### Macrovascular complications

No statistically significant differences in the risk of the composite macrovascular outcome were observed among ethnic minority groups compared to Dutch-origin.

#### Coronary outcomes

Ethnic differences in coronary outcomes showed clear heterogeneity across groups. The risk of angina pectoris was highest among Turks (HR 1.62 [95%-CI 1.26; 2.08]), followed by Dutch Caribbean, non-Europeans and Europeans. Similarly, myocardial infarction risk was increased among Turks (HR 1.69 [95%-CI 1.30; 2.21]) and Surinamese, while excess risk in Europeans was attenuated in fully adjusted models. Ischemic heart disease risk was elevated among Surinamese (HR 1.51 [95%-CI 1.10; 2.07]), whereas other groups showed no consistent differences.

Across coronary outcomes, environmental exposures consistently contributed to risk differences, mediating between 15 and 50% of observed associations. Key contributors included air pollution, environmental temperature, and neighborhood accessibility.

#### Cerebrovascular disease

Dutch Caribbean (HR 1.60 [95%-CI 1.17; 2.18]) and Surinamese (HR 1.45 [95%-CI 1.23; 1.71]) had higher risks of cerebral vascular accident, whereas no statistically significant differences were observed in other groups. These associations were primarily mediated by SES (32-33%), followed by environmental exposures (15-22%) and healthcare use (18-19%).

#### Peripheral artery disease

In contrast to other macrovascular outcomes, several ethnic minority populations showed lower risks of peripheral artery disease, including Indonesians, Moroccans, Surinamese, Turks and non-Europeans (e.g. HR 0.32 [95%-CI 0.20; 0.51] for Moroccans). This inverse association was partly mediated by (up to 42%) lifestyle factors, SES and baseline health. No statistically significant differences were observed for Dutch Caribbeans and Europeans.

#### Heart failure

Increased risk of heart failure was observed among Surinamese, Turks, and Europeans (e.g. HR 1.46 [95%-CI 1.19; 1.80] for Turks). Differences were most strongly explained by health care use (max 15%), environmental exposures (max 39%) and baseline health (max 12%).

#### Transient ischaemic attack

Surinamese had an increased risk of transient ischaemic attack (HR 1.39 [95%-CI 1.21; 1.72]; of which 10% was mediated by environmental factors. In contrast, lower risks were observed among Moroccans, Indonesians and non-Europeans (e.g. HR 0.51 [95%-CI 0.34; 0.76] for Moroccans), partially mediated by lifestyle factors (max 14%).

### Microvascular complications

Surinamese and Dutch Caribbeans were at increased risk of microvascular complications (e.g. HR 1.43 [95%-CI 1.20; 1.70] for Dutch Caribbeans), whereas no statistically significant differences were observed in other groups. Nephropathy risk was elevated among Indonesians, Surinamese and Dutch Caribbeans. Neuropathy was more common among Surinamese, Dutch Caribbean and non-Europeans, while Indonesians showed a lower risk of neuropathy (HR 0.71 [95%-CI 0.51; 0.99]). Retinopathy risk was consistently elevated across all ethnic minority groups compared with Dutch-origin, with the exception of Europeans (e.g. HR 1.87 [95%-CI 1.62; 2.17] for Moroccans).

Across microvascular outcomes, environmental exposures were the most influential mediating domain, followed by socioeconomic factors. Key environmental contributors included air pollution, population density, land-use mix, walkability, and neighbourhood accessibility indicators. Household composition, household type, income-related benefits and home ownership primarily drove SES mediation.

### Depression

The risk of depression was higher among Moroccans, Surinamese, Turks (e.g. HR 2.09 [95%-CI 1.75; 2.50]) and non-Europeans, whereas no statistically significant differences were observed for Indonesians, Dutch Caribbeans or Europeans.

Environmental exposures were the strongest mediating domain, followed by SES, baseline health indicators and medication use. Key environmental factors included air pollution, neighborhood deprivation, food environment and built environment characteristics. SES mediation was driven by household composition and income-related benefits, while baseline health indicators (including comorbidity burden, blood pressure and sleep-related symptoms) and medication use (analgesics) contributed to a lesser extent.

### Additional analyses

Second-generation migrants were younger than first-generation migrants and had lower absolute prevalences of complications. Stratified by migration generation were exploratory (Supplementary File 7). Risk patterns were broadly similar across generations, with slightly higher risks among second-generation Indonesians and non-Europeans relative to first-generation individuals, with Dutch-origin as reference group.

## Discussion

In this large, nationally representative cohort of adults with T2D in the Netherlands, we found substantial but heterogeneous ethnic differences in risks of diabetes-related complications and mortality. These differences varied markedly by outcome and ethnic group and were partially attenuated after accounting for environmental exposures, SES, lifestyle and baseline health indicators. Together, these findings support the multifactorial nature of ethnic differences in T2D outcomes and highlight the importance of context-specific approaches to prevention and care. It should be noted that the reported differences are expressed as relative risks; absolute risk differences may be modest for several outcomes, and should be considered when interpreting clinical relevance.

Surinamese individuals consistently showed elevated risks for multiple T2D complications, with all-cause mortality being the exception. These findings are consistent with a meta-analysis on ethnic differences in T2D complications in Europe [3], and may reflect a relatively lower cancer mortality among Surinamese compared to Dutch [31]. However, the Surinamese population is heterogeneous, and previous studies have shown that individuals of South Asian (Hindustani) origin have a substantially higher risk of T2D and related complications compared with those of African (Creole) origin [2]. As our data did not allow us to distinguish between these subgroups, potential differences within the Surinamese population could not be explored. The high contribution of environmental exposures, including air pollution, urban heat exposure and population density, to the attenuation of risk estimates suggests that neighborhood-level contextual factors may contribute substantially to cardiometabolic risk in this group. This supports the growing evidence that environmental and contextual determinants contribute substantially to ethnic inequalities in T2D risk [32]. Baseline health indicators such as multimorbidity, hypertension and higher BMI further contributed to excess risk. The combination of environmental and clinical vulnerability may help explain the broad pattern of elevated risks, although causal inference could not be established.

Turks and Europeans were at elevated risk for several macrovascular outcomes compared to Dutch-origin, including angina pectoris and myocardial infarction. This increased risk among Turks aligns with a study by Armengol et al., reporting up to six times higher odds for coronary heart disease compared to Dutch [6]. Mediation analyses indicated that environmental factors were the most important mediating domain, explaining approximately 10-17% of the excess risk for angina pectoris. Key contributors included air pollution, environmental temperature, and indicators of neighbourhood accessibility such as road travel time and land-use density. Although these proportions appear modest, they reflect the potential impact of widely distributed environmental exposures that affect entire populations. While genetic factors may also play a role [33], these findings point to the relevance of environmental and healthcare-related determinants that are amenable to intervention.

Moroccans and Indonesians generally showed lower risks of several macrovascular complications (e.g. peripheral artery disease, transient ischemic attack). For Moroccans, smoking was the dominant mediator, indicating that lower smoking prevalence may provide substantial cardiometabolic protection. Medication use helped explain their lower CVD mortality, although the mechanisms (e.g. treatment adherence, prescribing patterns or care-seeking) cannot be disentangled with available data [34]. For Indonesians, medication use and healthcare use contributed modestly to the observed patterns. These findings reflect treatment- and care-related processes, rather than lifestyle behaviors. Taken together, these results show that favorable behavioral or care-related factors may mitigate risk in certain ethnic groups, underscoring the non-uniform nature of ethnic differences.

Risks of microvascular disease were elevated among Surinamese and Dutch Caribbean individuals, and retinopathy risk was elevated in all minority groups except Europeans, which is in line with previous findings [9, 35]. Environmental exposures were consistently identified as the most important mediating domains in the attenuating ethnic differences, followed by socioeconomic factors.

Depression risks were higher among Moroccans, Surinamese, Turks and non-Europeans. Environmental exposures were the strongest mediating domain, followed by baseline health status and healthcare-related factors, including pain management, jointly influence mental health outcomes. These patterns may also reflect the role of psychosocial stressors, including experiences of discrimination, social marginalization and acculturation challenges, which have been linked to both depression and adverse diabetes outcomes in ethnic minority populations [36, 37].

Our findings are consistent with prior multi-ethnic studies in the Netherlands and the UK [7], but extend the literature by investigating the most influential mediators from a large candidate set including environmental and healthcare-related ones. The strong influence of environmental exposures supports emerging evidence that upstream determinants are central to shaping T2D outcomes [19], particularly in ethnically diverse populations [38]. In addition, socio-economic status and healthcare use also contributed substantially to the observed differences, whereas lifestyle factors and medication use played a more limited role. Since most aspects of our environment are modifiable, this provides considerable potential for population-level prevention.

### Strengths and limitations

This study has several strengths and limitations. First, residual confounding is possible due to unmeasured factors such as sleep, genetic variation, cultural practices and psychosocial stress. Baseline HbA1c required multiple imputation, introducing some uncertainty. Sample sizes were limited for several subgroups, especially Dutch Caribbeans and second-generation migrants, reducing precision. Exclusion of specialist-treated patients may have underestimated risks if severe cases, potentially overrepresented among ethnic minority populations, were missed. In the variable-selection step, Shapley-based interpretation identified predictors such as influenza vaccination, distance to daycare, and environmental temperature. These likely act as proxies for underlying health status, urbanicity, or broader environmental context rather than causal determinants. The data-driven approach used in this study identifies predictors based on their contribution to prediction rather than causal relationships. Therefore, findings should be interpreted as indicative of predictive relevance rather than causal priority. Finally, the large number of outcomes and subgroups raises the possibility of multiple testing. Therefore, we interpreted findings based on consistency across models. Strengths of this work include the large, nationally representative cohort, linkage of primary care data with high-quality administrative registries and robust outcome ascertainment. The use of machine-learning to select explanatory variables enhanced the relevance and precision of the mediation analysis, by enabling the identification of the most influential predictors from a large set of candidate factors, while accounting for complex interactions between variables.

### Implications

Our findings highlight substantial heterogeneity in diabetes-related complications across ethnic groups in the Netherlands, underscoring the need for tailored approaches to prevention and care that take into account environmental and socio-demographic context. In particular, the observed variation in complication patterns shows that risks differ across outcomes and ethnic groups. Clinicians should therefore not assume uniformly elevated risk, but instead be alert to specific complications that are more common in certain groups.

Environmental exposures emerged as consistent mediators across multiple outcomes. Their ubiquity and modifiability suggests that they are relevant targets for population-level prevention strategies, particularly through urban planning, air quality regulation and interventions targeting neighbourhood liveability.

Our findings also suggest that differences in healthcare utilization and baseline health status may contribute to observed disparities, indicating that more equitable access to preventive and ongoing care could help reduce inequities in outcomes.

Finally, the observed heterogeneity within broad ethnic categories emphasizes the importance of moving beyond aggregated groupings in both research and policy, as aggregated categories may obscure clinically and socially meaningful variation in risk profiles.

## Conclusion

In conclusion, ethnic differences in T2D complications and mortality in the Netherlands are substantial, multifactorial and outcome-specific. This study shows that these disparities are only partly explained by clinical factors and are substantially shaped by contextual and environmental circumstances. These findings highlight the importance of addressing contextual and system-level drivers of T2D complication risk, while accounting for heterogeneity within and between ethnic groups.

## Author contributions

MM contributed to funding acquisition, study conception and design, conducted all statistical analyses, drafted the manuscript including figures and tables, and is the guarantor of the work. BTS contributed to the analytic workflow, accessed and verified data in the environment of Statistics Netherlands and contributed to interpretation of results and critical revision of the manuscript. PJEM, FR, GN, JAO, RMCH, MTB, JWJB contributed to the design and set-up of the DIAMANT study, facilitated data acquisition and contributed to interpretation of the findings and critical revision of the manuscript. JWJB also contributed to study supervision and funding acquisition. IV and JL contributed expertise related to the GECCO data and critically revised the manuscript, JL also contributed to GECCO data acquisition. All authors contributed to revising the manuscript and approved the final version.

## Declaration of generative AI and AI-assisted technologies in the writing process

During the preparation of this work the authors used Microsoft 365 Copilot in order to improve readability and language. After using this tool, the authors reviewed and edited the content as needed and take full responsibility for the content of the publication.

## Data availability

The dataset used for this study is not publicly available, because it cannot leave the environment of Statistics Netherlands according to their privacy regulations. Requests to access the dataset should be directed to m.muilwijk@amsterdamumc.n; authorization from Statistics Netherlands is required.

## Acknowledgements

The authors would like to thank all the healthcare providers contributing information to the PHARMO Data Network.

